# Detection and removal of pacing artifacts prior to automated analysis of 12-lead ECG

**DOI:** 10.1101/2021.01.21.21250278

**Authors:** Kazi T. Haq, Neeraj Javadekar, Larisa G. Tereshchenko

## Abstract

**Background:** Pacing artifacts must be excluded from the analysis of paced ECG waveform. This study aimed to develop and validate an algorithm to identify and remove the pacing artifacts on ECG.

**Methods:** We developed a semi-automatic algorithm that identifies the onset and offset of a pacing artifact based on the ECG signal’ slope steepness and designed a graphical user interface that permits quality control and fine-tuning the constraining threshold values. We used 1,054 ECGs from the retrospective, multicenter cohort study “Global Electrical Heterogeneity and Clinical Outcomes,” including 3,825 atrial and 10,031 ventricular pacing artifacts for the algorithm development and 22 ECGs including 108 atrial and 241 ventricular pacing artifacts for validation. Validation was performed per digital sample. We used the kappa-statistic of interrater agreement between manually labeled sample (ground-truth) and automated detection.

**Results:** The constraining parameter values were for onset threshold 13.06±6.21 μV/ms, offset threshold 34.77±17.80 μV/ms, and maximum window size 27.23 ± 3.53 ms. The automated algorithm detected a digital sample belonging to pacing artifact with a sensitivity of 74.5% and specificity of 99.6% and classified correctly 98.8% of digital samples (ROC AUC 0.871; 95%CI 0.853-0.878). The kappa-statistic was 0.785, indicating substantial agreement. The agreement was on 98.81% digital samples, significantly (*P*<0.00001) larger than the random agreement on 94.43% of digital samples.

**Conclusions:** The semi-automated algorithm can detect and remove ECG pacing artifacts with high accuracy and provide a user-friendly interface for quality control.

**Highlights:**

- We developed and validated a semi-automated algorithm to detect and remove pacing spike artifacts from a digital ECG signal.
- The semi-automated algorithm can detect and remove pacing spike artifacts with high accuracy and provide a user-friendly interface for quality control.

## Introduction

Since the 1950s, cardiac pacing became a vital treatment modality for a growing number of cardiovascular patients. Innovations in cardiac pacing expanded indications for implantable devices capable of delivering cardiac pacing.[1] Pacemaker implantation rates increased from 467 per million in 1993 to 616 per million in 2009.[2] In 2014, an estimated 351,000 pacemaker inpatient procedures were performed in the US.[2] The number of patients living with an implanted cardiac pacemaker is steadily growing.

An electrocardiogram (ECG) is widely used to determine the heart rhythm and to evaluate the performance of pacemaker functioning, especially in emergency settings.[3] Atrial-paced and ventricular-paced rhythm and atrioventricular (AV) dual-paced rhythm are included in the list of core primary ECG diagnostic statements, endorsed by the American Heart Association (AHA), the American College of Cardiology (ACC), the Heart Rhythm Society (HRS), and the International Society for Computerized Electrocardiography (ISCE).[4]

Notably, the ECG diagnostic standards enforced the rule that no secondary statements can accompany the primary diagnostic statement of paced rhythm or paced complexes.[4] The rule[4] stemmed from the dogma about secondary repolarization abnormalities, stating that paced ventricular complexes are examples of secondary repolarization abnormalities.[5] Therefore, the consensus is that paced ECG could only be used to diagnose paced rhythm, but, otherwise, it cannot be clinically useful.[5]

Nevertheless, we,[6] and others, showed that cardiac memory could be detected during continued altered activation.[7] Cardiac memory is neither pure primary nor pure secondary repolarization abnormality.[8, 9] Spatial ventricular gradient (SVG) is independent of the activation sequence.[6, 10, 11] Measurement of SVG on paced ECG furnished clinically useful information.[12, 13] Thus, paced ECG carries important data for meaningful analysis, which needs to be further studied.

The first step in the automated analysis of paced ECG is the removal of pacing artifacts. No such algorithm has been previously developed that can both detect and remove a pacing spike artifact from the 12-lead and orthogonal XYZ ECG. This paper presents the development and validation of a novel semi-automated algorithm to detect and remove pacing artifacts. We tested the hypothesis that the newly developed semi-automated algorithm can accurately detect and remove pacing artifacts from a vectorcardiogram (VCG) obtained from 12-lead ECG.

## Methods

### Study population

We analyzed data from the retrospective, multicenter cohort study “Global Electrical Heterogeneity and Clinical Outcomes” (GEHCO).[13, 14] The study was approved by the Institutional Review Boards at the Oregon Health & Science University and each participating institution. The study collected digital 12-lead ECG signal recorded before implantation of implantable cardioverter-defibrillator (ICD) or cardiac resynchronization therapy defibrillator (CRT-D). The present study included only patients with atrial-paced (AP), ventricular-(including bi-ventricular-) paced (VP), and AV dual-(including atrial and bi-ventricular) paced (AVP) rhythm on available digital 12-lead ECG. Only one (pre-implant) ECG per patient was included in this study.

### Algorithm description

The algorithm and open-source software code written in MATLAB (MathWorks, Natick, MA, USA) are provided at https://github.com/Tereshchenkolab/Pacing_spike_removal. The algorithm was developed using 10-second digital ECG signal with sampling rate 500Hz. The amplitude resolution was either 1 µV or 5 µV.

Figure 1 shows the flowchart of the developed algorithm. First, baseline wander was removed from the 12-lead ECG. Then XYZ orthogonal ECG was obtained from a 12-lead ECG signal by using the Kors transformation matrix.[15] Once the XYZ leads were obtained, the vector magnitude was calculated from Eq.1.

**Figure 1.**
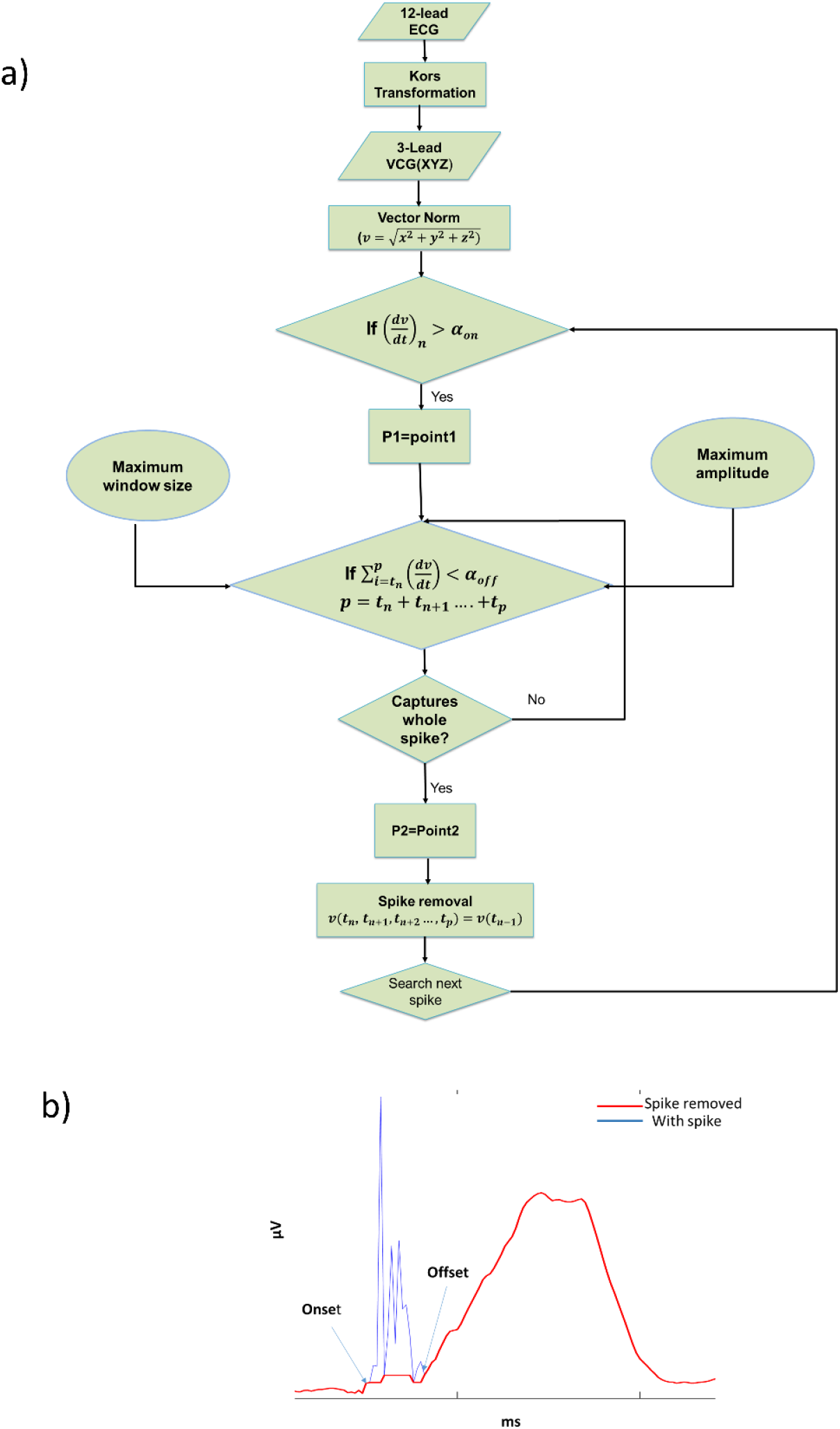
A. Flowchart representation of the algorithm to detect and remove a pacing artifact. B. Representative example of a pacing artifact on vector magnitude ECG signal.

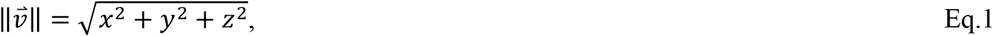

where 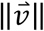is the vector norm, and x, y, and z are the XYZ orthogonal vectors.

#### Step 1. Pacing artifact onset detection

Slope 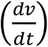 was calculated for each pair of the consecutive samples on the 10-second ECG recording. The algorithm automatically selected the pacing artifact’s onset (*P*_*1*_) if the following condition was satisfied.

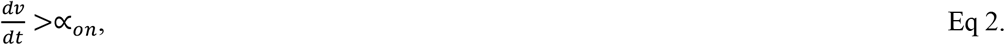

where α_on_ = pacing artifact onset threshold.

#### Step 2. Pacing artifact offset detection

Next, the algorithm searched for the pacing artifact’s offset. Since the vector norm is an absolute number, the sum of the slopes measured for each pair of the consecutive samples within the pacing artifact tends to zero, assuming that the pacing artifact’s offset approaches the baseline, which has a value close to zero. However, in practice, the pacing artifact’s offset has a non-zero value due to the saturation effect and baseline deviation. Therefore, we assumed that the offset was an arbitrary positive value, which we referred to as pacing artifact offset threshold, α_off._ Thus, the offset point (*P*_*2*_) was obtained when the following condition was satisfied.

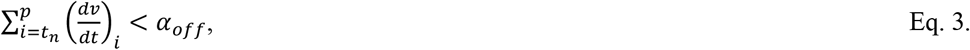

where *p*=*t*_*n*_+*t*_*n*+1_ … … … … … … .+*t*_*p*_and α_off_ = pacing artifact’s offset threshold.

#### Step 3. Pacing artifact removal

Once the onset (*P*_*1*_) and offset (*P*_*2*_) time points of a pacing artifact were determined, the spike was removed by making the signal values within the detected pacing artifact time window (*P*_*1*_ *– P*_*2*_) equal to the value at the adjacent time point just before *P*_*1*._ This was given by,

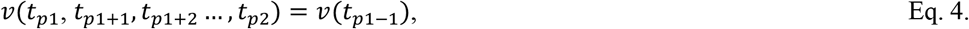

where *v* = signal amplitude.

#### Graphic user interface

A graphic user interface was developed that allows users to choose the threshold values from a given range (Figure 2). Additional user-defined parameters were included. Maximum amplitude defines the range of the upper limit value of the signal. Maximum window size defines the range of the sample points considered as the time window of the pacing spike. Re-run allows re-run of the algorithm recursively if a satisfactory result cannot be obtained by altering the thresholds several times. As shown in Figure 2, the dv/dt curve (orange) is superimposed on the vector magnitude plot, which helps the user check whether the maximum dv/dt falls within the QRS complex window.

**Figure 2.**
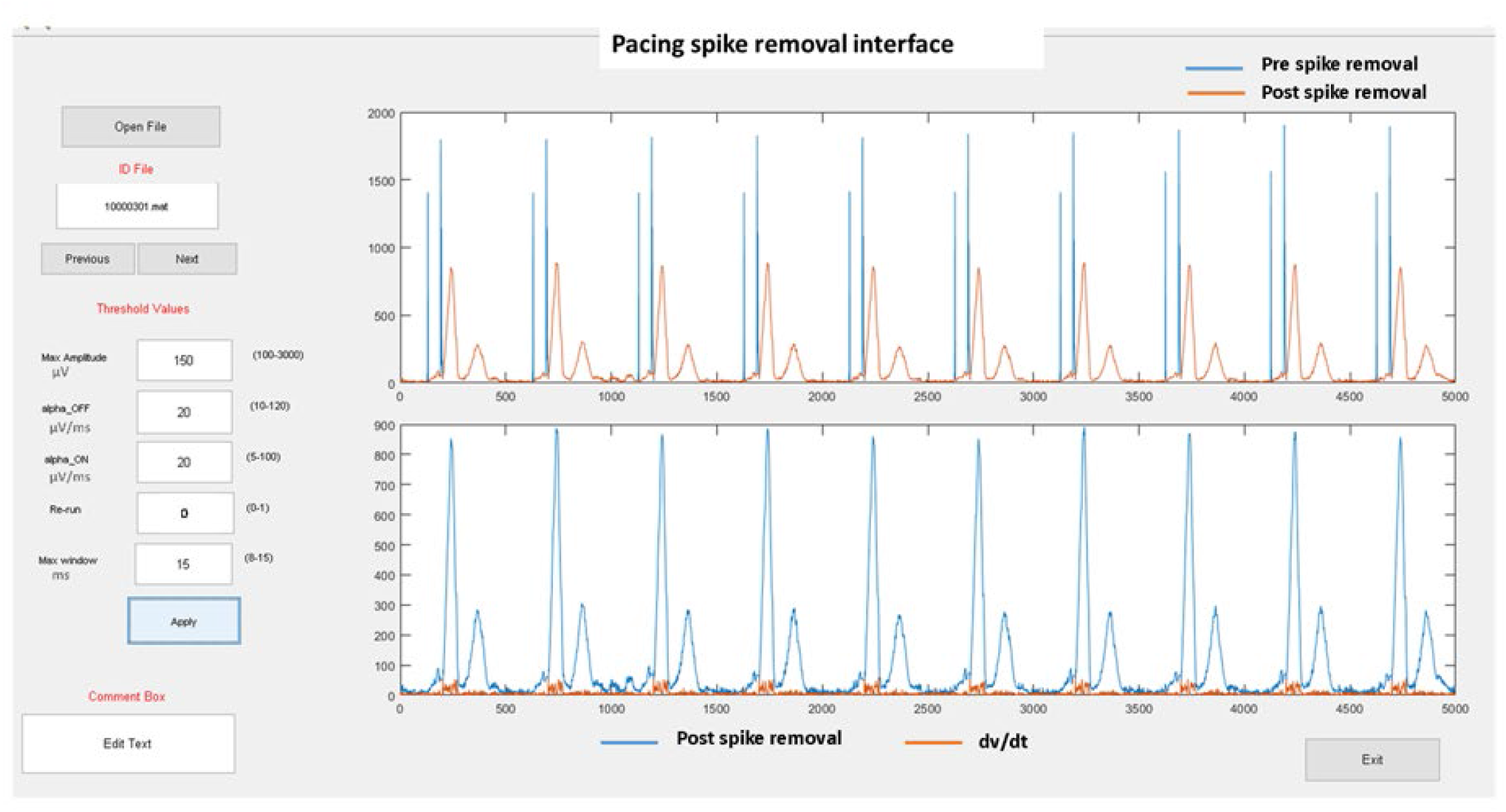
Graphic user interface for pacing artifact removal. The interface allows users to input threshold parameters and review the output for each set of chosen parameters.

### Validation of the automated pacing artifacts detection

Considering the broad differences in pacing artifacts morphology and duration (because of differences in ECG recording equipment and implanted device manufacturer), we used 98% of the data (1,054 ECGs including 1,399 atrial, 7,605 ventricular, and 2,426 AV pacing artifacts) for the algorithm development and 2% of the data (22 ECGs including 108 atrial and 241 ventricular pacing artifacts) for validation.

For validation, to obtain the ground truth, one investigator (NJ) manually labeled each sample on digital ECG signal as either belonging to pacing artifact (Yes) or not (No). Another investigator (KTH) ran an automated algorithm and similarly reported each data sample results as pacing artifact Yes or No. The investigators (NJ and KTH) were blinded to each other results. The third investigator (LGT) conducted statistical analysis.

### Statistical analysis

Statistical analysis was performed using STATA MP 16.1 (StataCorp LLC, College Station, TX, USA). Continuous variables were reported as mean ± standard deviation (SD). We used the kappa-statistic measure of interrater agreement for two independent raters. Nonparametric receiver operating characteristic (ROC) analysis with a rating and discrete classification data was performed to calculate the area under the ROC curve (ROC AUC) and measure the automated pacing artifact detection’s sensitivity and specificity.

## Results

### Study population

Clinical and demographic characteristics of the study population are reported in Table 1. This study included heart failure patients, mostly white men. Approximately half of the patients had nonischemic cardiomyopathy, and one-third had diabetes. Most of the patients (70%) had CRT-D implanted, and 30% had ICD implanted. Implanted devices were manufactured by four companies (Table 1), which allowed us to develop the algorithm that considered various features of pacing pulses by different manufacturers. The vast majority of patients (86%) had a VP or AVP rhythm on analyzed ECG. An average heart rate was 70 beats per minute.

**Table 1.**
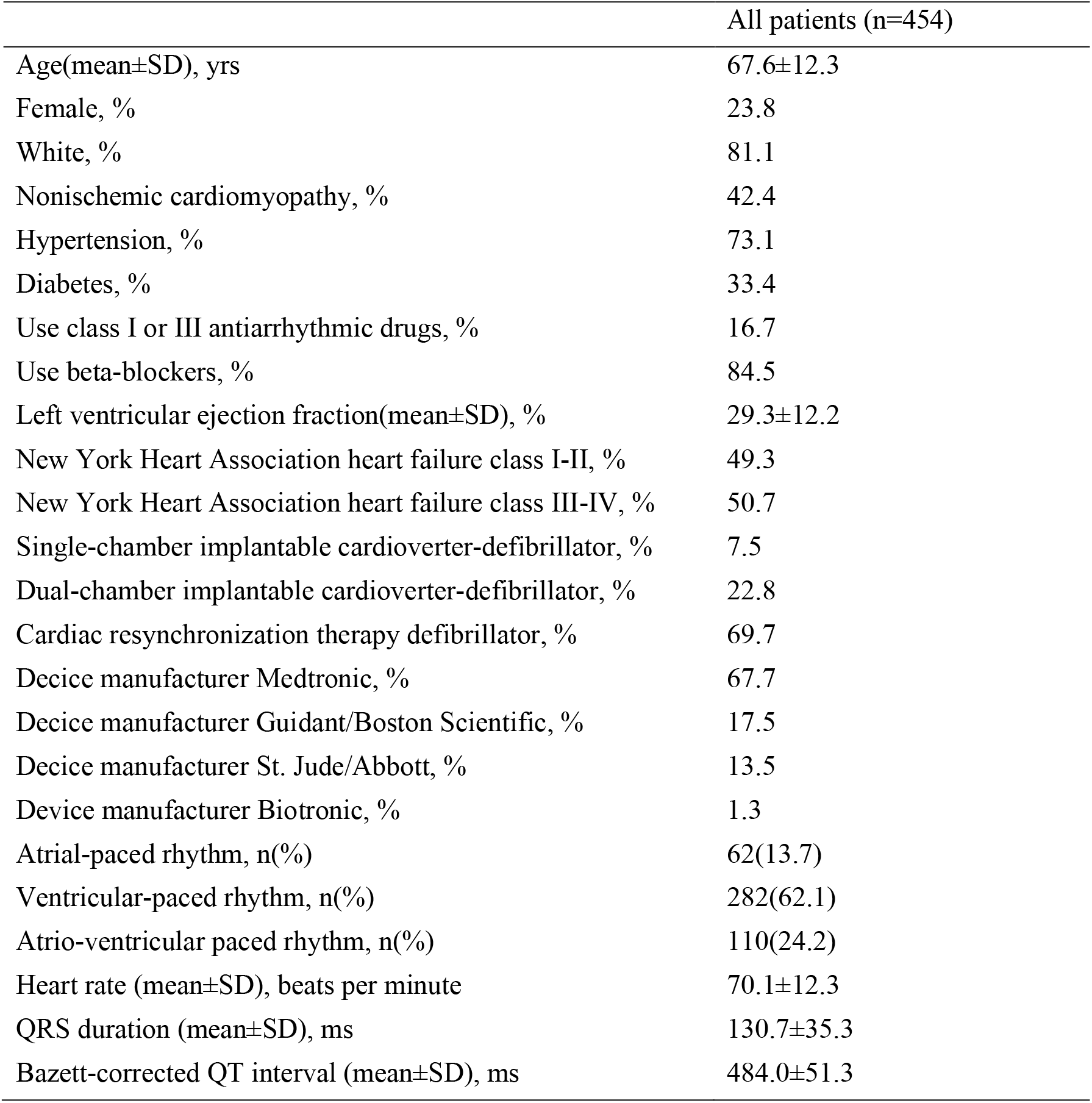
Study population characteristics.

|  | All patients (n=454) |
| --- | --- |
| Age(mean±SD), yrs | 67.6±12.3 |
| Female, % | 23.8 |
| White, % | 81.1 |
| Nonischemic cardiomyopathy, % | 42.4 |
| Hypertension, % | 73.1 |
| Diabetes, % | 33.4 |
| Use class I or III antiarrhythmic drugs, % | 16.7 |
| Use beta-blockers, % | 84.5 |
| Left ventricular ejection fraction(mean±SD), % | 29.3±12.2 |
| New York Heart Association heart failure class I-II, % | 49.3 |
| New York Heart Association heart failure class III-IV, % | 50.7 |
| Single-chamber implantable cardioverter-defibrillator, % | 7.5 |
| Dual-chamber implantable cardioverter-defibrillator, % | 22.8 |
| Cardiac resynchronization therapy defibrillator, % | 69.7 |
| Decice manufacturer Medtronic, % | 67.7 |
| Decice manufacturer Guidant/Boston Scientific, % | 17.5 |
| Decice manufacturer St. Jude/Abbott, % | 13.5 |
| Device manufacturer Biotronic, % | 1.3 |
| Atrial-paced rhythm, n(%) | 62(13.7) |
| Ventricular-paced rhythm, n(%) | 282(62.1) |
| Atrio-ventricular paced rhythm, n(%) | 110(24.2) |
| Heart rate (mean±SD), beats per minute | 70.1±12.3 |
| QRS duration (mean±SD), ms | 130.7±35.3 |
| Bazett-corrected QT interval (mean±SD), ms | 484.0±51.3 |

### Algorithm development and validation

For the algorithm development, to determine the *P*_*1*_ and *P*_*2*_ thresholds, we detected and removed 1,399 atrial, 7,605 ventricular, and 2,426 AV pacing artifacts. Figure 3 shows a representative example of pacing artifacts removal from a VP VCG vector magnitude signal. Figure 4 shows another example where pacing artifacts were removed from an AVP VCG vector magnitude signal.

**Figure 3.**
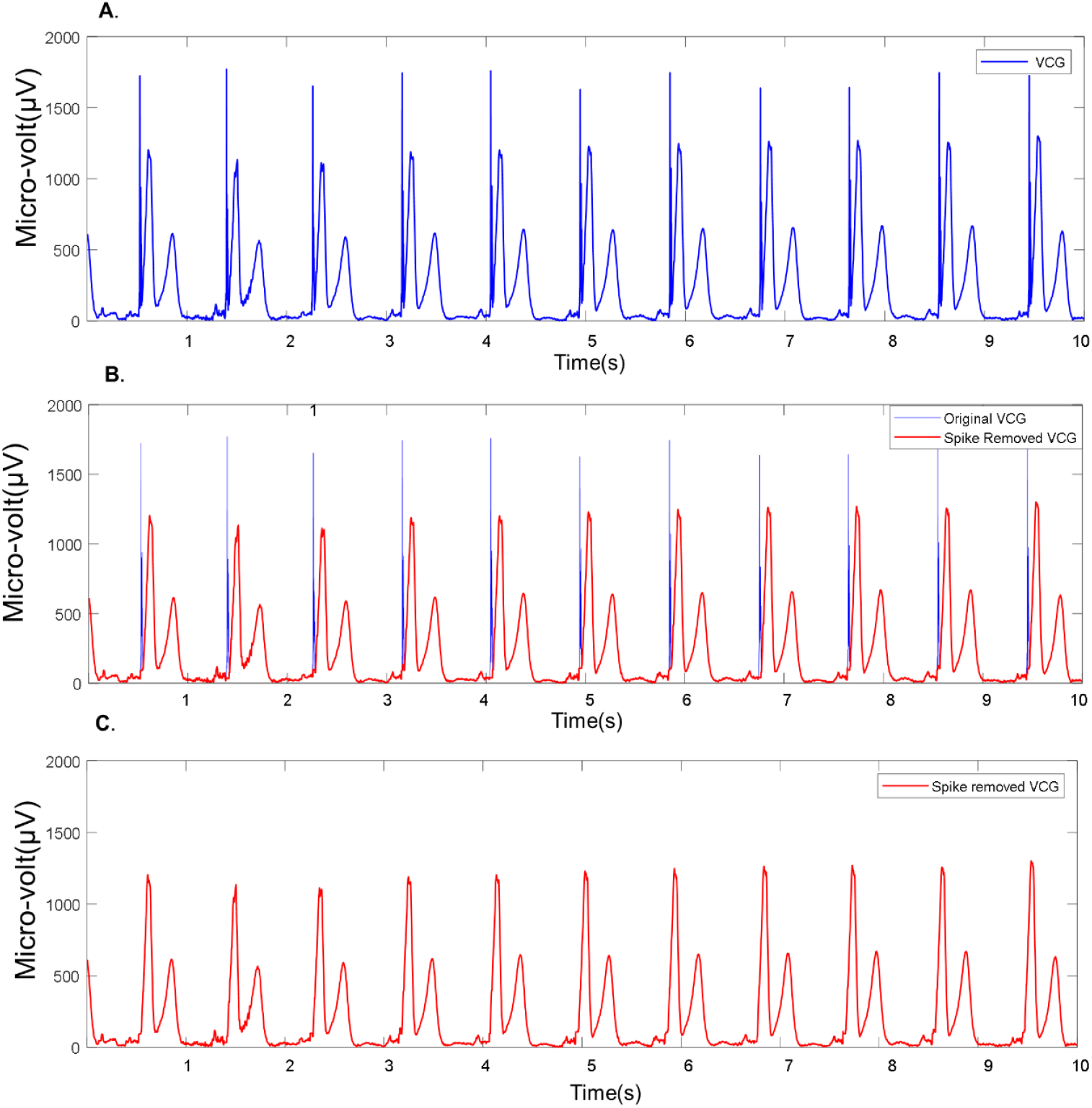
Example of pacing artifact removal from a ventricular-paced VCG. A) Original ventricular-paced VCG vector magnitude signal. B) Detected pacing artifacts (blue) and VCG signal (red). C) VCG vector magnitude signal after removal of pacing artifacts. The pacing spike onset (*α*_*on*_) and offset (*α*_*off*_)threshold values were 5 and 10 µV/ms, respectively.

**Figure 4.**
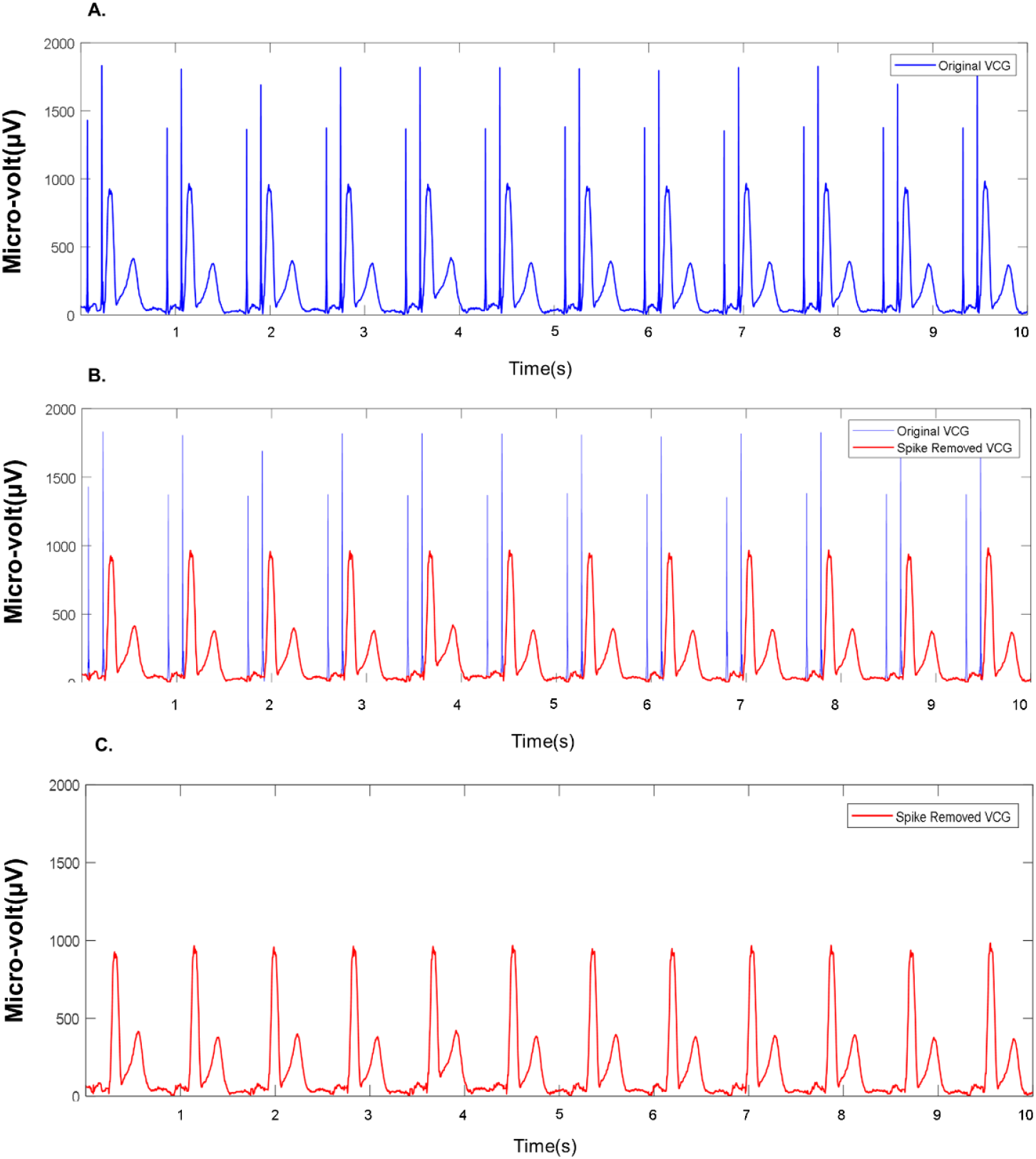
Example of pacing spike removal from AV-paced VCG. A) Original AV-paced VCG vector magnitude signal. B) Detected pacing artifacts (blue) and VCG signal (red). C) VCG vector magnitude signal after removal of pacing artifacts. The corresponding value of *α*_*on*_ and *α*_*off*_ were 7.5 and 15.5 µV/ms, respectively.

**Figure 5.**
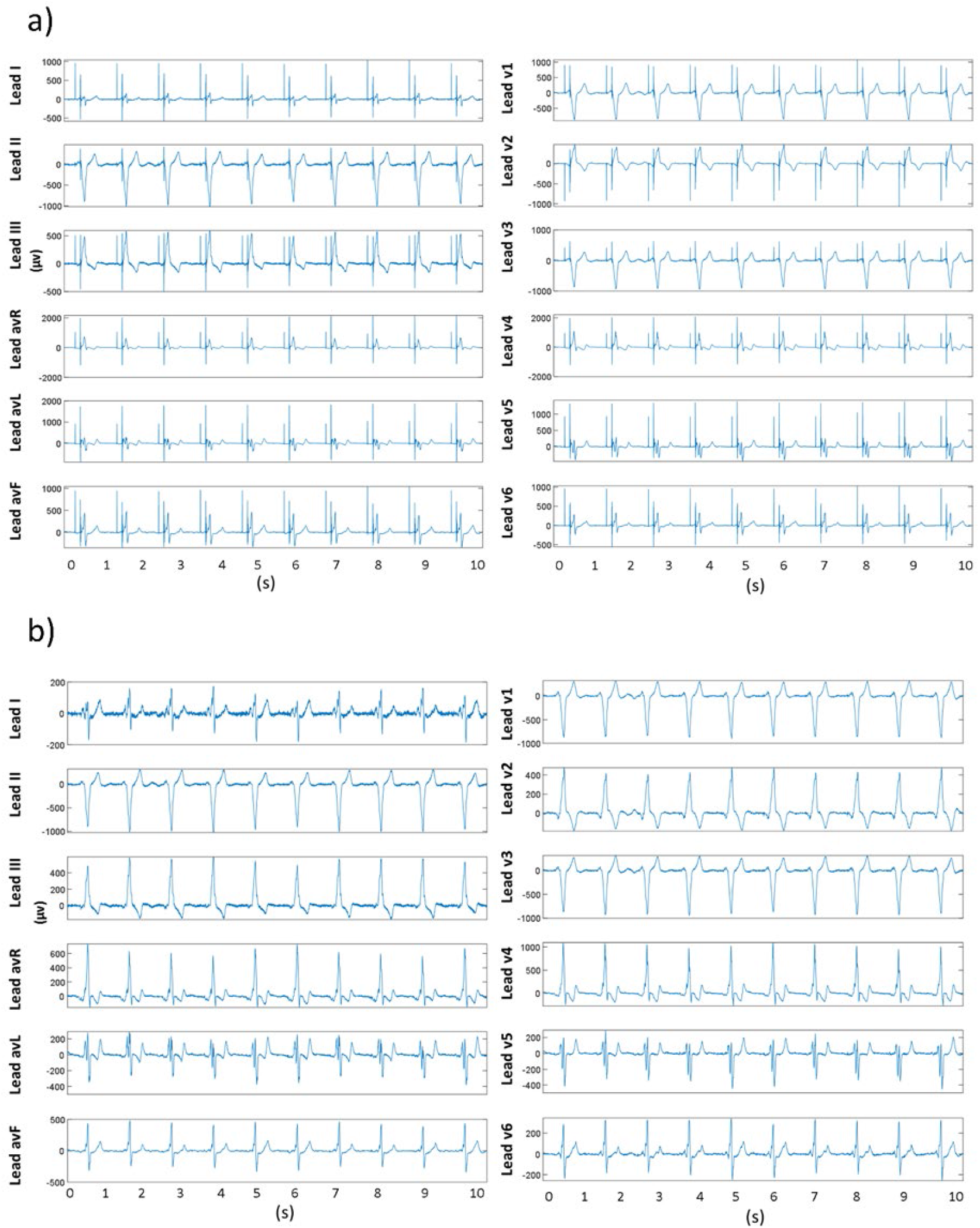
Example of pacing artifacts removal from 12-lead ECG a) 12-lead ECG with pacing artifacts. b) 12-lead ECG trace after pacing artifacts removal.

From the validation dataset (n= 22 ECGs) the mean of 4 constraining parameter values were found as : mean maximum amplitude 146.36±76.36 μV, mean onset threshold (α_on_) 13.06±6.21 μV/ms, mean offset threshold (α_off_) 34.77±17.80 μV/ms, and mean maximum window size 27.23 ± 3.53 ms.

For validation of the automated algorithm, we analyzed 110,000 digital signal samples. Each ECG had 5000 samples. On average, a pacing artifact occupied 9.2±3.2 samples or 18.4±6.4 ms. Notably, the automated algorithm detected the presence of pacing artifact with 100% accuracy, 100% sensitivity, and 100% specificity.

Two-by-two table (Table 2) reports an agreement between the ground truth and automated detection of pacing artifacts for each sample of the digital ECG data. If the automated algorithm had made the determination randomly (but with probabilities equal to the overall proportions), we would expect the agreement on 94.43% of digital samples. In fact, they agreed on 98.81%. The amount of agreement indicated that we could reject the null hypothesis that they were making their determination randomly (*P*<0.00001). The kappa-statistic was 0.785, indicating substantial agreement.

**Table 2.**
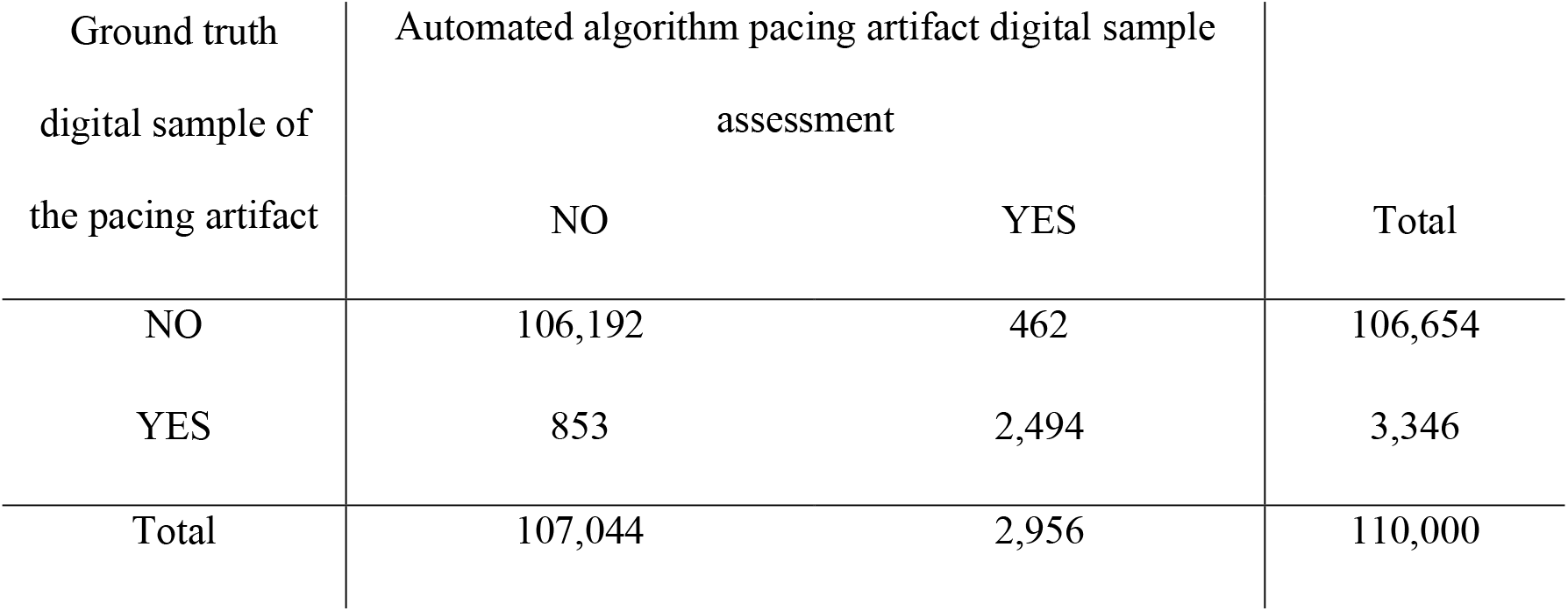
Two by two table of a per-sample agreement between the ground trith and automated algorithm detection of pacing artifacts.

| Ground truth<br>digital sample of<br>the pacing artifact | Automated algorithm pacing artifact digital sample<br>assessment |  | Total |
| --- | --- | --- | --- |
|  | NO | YES |  |
| NO | 106,192 | 462 | 106,654 |
| YES | 853 | 2,494 | 3,346 |
| Total | 107,044 | 2,956 | 110,000 |

The automated algorithm assigned a digital sample to pacing artifact with ROC AUC 0.871 (95% confidence interval 0.853-0.878), a sensitivity of 74.5%, and specificity of 99.6% and classified correctly 98.8% of digital samples.

## Discussion

In this work, we developed and validated the semi-automated algorithm to detect and remove pacing artifacts from a digital ECG signal. The fully automated algorithm was perfectly 100% accurate in detection of a pacing artifact’s presence, and demonstrated 75% sensitivity and 100% specificity for the per-digital-sample automated detection of pacing artifacts, whereas a user-friendly interface allowed additional fine-tuning and quality control of the artifact removal.

Current ECG machines convert the analog ECG signal to digital at the front end.[16] Modern pacemaker’s stimulus output is frequently ≤ 0.25 ms in duration. Therefore, front-end sampling has to be ≥ 10,000 samples per second in order to detect and represent the pacemaker’s stimulus output. Furthermore, contemporary bipolar pacemaker’s stimulus output is usually small (2-4 Volt). The AHA/ACC/HRS/ISCE-endorsed recommendations for the standardization and interpretation of the ECG[16] emphasized that ECG manufacturers should maintain the required front-end sampling rate for reliable and accurate detection of narrow pacemaker pulses but should not artificially increase pulses’ amplitude to avoid ECG’s morphology distortion.

However, in clinical settings, the information about the front-end sampling rate and an approach to handling the presentation of pacemaker stimulus outputs by a specific ECG recording machine is not readily available. In a nonselective CRT patient population, cardiologists observed visible ventricular pacing artifacts in 90% and visible atrial pacing artifacts – in 70% of patients with paced rhythm, whereas the automated ECG reading algorithm detected only 20% of patients with VP rhythm, and none with AP rhythm.[17]

In accordance with the growing population of patients with implanted pacemakers, ICD, and CRT devices, the list of meaningful interpretations of paced ECG morphology is expanding.

Acute myocardial infarction can manifest on ECG in patients with VP rhythm, although the sensitivity of acute myocardial infarction diagnosis on paced ECG is low.[18, 19] Left ventricular paced QRS width and the difference between biventricular-paced and pre-implant QRS width predict CRT response.[20] Our previous study showed that the addition of global electrical heterogeneity (GEH) ECG metrics to clinical risk factors of sudden cardiac death (SCD) is especially rewarding in the presence of paced rhythms.[12] In a subgroup of participants with VP ECG (Supplemental Table 13 in [12]), with the addition of GEH parameters, 33% of SCD victims were appropriately reclassified into a higher-risk category (from low to high risk). In contrast, only 10% were similarly appropriately reclassified amongst participants without a paced rhythm on ECG. Furthermore, in the VP rhythm subgroup, no SCD victims were inappropriately reclassified from high to low risk. The addition of GEH also improved SCD-specific risk prediction. The proportion of SCD decreased from 12% to 6% in the intermediate-risk group and increased from 15% to 18% in the high-risk group. The large prospective study of more than 20,000 participants with a median of 14 years of follow-up showed clinical usefulness of GEH measurements on VP ECG.[12]

Therefore, there is a growing need for reliable detection of pacing artifacts and accurate analysis of paced ECG morphology. Particularly, while the pacing artifact detection is important for the diagnosis of a paced rhythm, the pacing artifact itself should be removed from the analysis of the paced ECG waveform. As discussed above, an ideal solution endorsed by the AHA/ACC/HRS/ISCE involves the oversampling at the ECG front end at ≥10,000 Hz, saving pacing artifacts data on a unique marker-channel, and subsequent downsampling to 500-1000Hz for conventional ECG signal analysis and storage.[16] For example, the algorithm described by Polpetta and Banelli[21] follows the recommended approach and reports promising results.

Unfortunately, in clinical settings, it is usually unknown which front-end algorithm is utilized by a given ECG machine manufacturer. If an ECG has been recorded using unknown front-end characteristics, signal processing is the only option. A few previous fully automated algorithms addressed pacing artifacts detection. Helfenbein et al.[22] developed an algorithm for pacing artifacts detection on a commonly used ECG signal (sampling rate 500Hz, bandwidth 0.05-150 Hz), capitalizing on the fact that low-pass filtering broadens the pacing artifact width. The authors reported a sensitivity of 97.2% for detecting a paced rhythm,[22] which is lower than our results. Notably, the authors did not report an accuracy and algorithm performance per each sample of the digital ECG signal. Furthermore, widely used algorithms[22] are based on filtering the ECG signal, which distorts the beginning of the QRS and widens the QRS complex. The distortion of the QRS complex challenges the clinical use of paced QRS morphology measurements.[20]

Up-to-date, our semi-automated algorithm is the only available solution for careful and accurate pacing artifacts removal, as required for the analysis of QRS morphology. Allowing the user to select the dynamic thresholds from a range of values offers the required flexibility, considering different pacing pulse characteristics produced by a wide range of pacemakers’ manufacturers. Moreover, the user can monitor spike removal’s effects on the signal and choose an alternative set of thresholds until satisfactory results are achieved.

### Limitations

This paper presented a semi-automated algorithm to detect and remove pacing spikes artifacts from ECG. However, the algorithm itself is not fully automatic. Semi-automated approach is time-consuming. However, the pacing spikes from diverse patient populations and pacemaker settings always come with a high degree of variability, making automated dynamic threshold estimation challenging, though not impossible. We have to emphasize that the ideal approach for the analysis of paced ECG waveform morphology has to include ECG front-end oversampling[16], recording the presence of pacing artifacts on a separate marker-channel, and immediate its removat at the front end. Removal of a pacing artifact at the front end preserves paced ECG morphology for its subsequent analysis. Growing number of patients with implanted pacemaker devices and clinical needs for a meaningful analysis of paced ECG waveform[12, 13, 20] calls for ECG manufacturers attention to handling ECG front-end manufacturing.

## Conclusions

We developed a semi-automated algorithm to detect and remove pacing spike artifacts from digital ECG signal. The algorithm demonstrated its ability to detect and remove pacing spike artifacts with high sensitivity and specificity.

## Data Availability

The algorithm, open-source software code written in MATLAB (MathWorks, Natick, MA, USA), and de-identified data example are provided at https://github.com/Tereshchenkolab/Pacing_spike_removal.

## Acknowledgment

The authors thank all GEHCO investigators.

